# Clinical Evidence for Improved Outcomes with Histamine Antagonists and Aspirin in 22,560 COVID-19 Patients

**DOI:** 10.1101/2021.03.29.21253914

**Authors:** Cameron Mura, Saskia Preissner, Susanne Nahles, Max Heiland, Philip E. Bourne, Robert Preissner

## Abstract

COVID-19 has spurred much interest in the therapeutic potential of repurposed drugs. A family of acid-reducing drugs, known as histamine H_2_ receptor antagonists (H2RA), competitively bind the H2R and block its stimulation by histamine; examples of such drugs are famotidine (e.g., Pepcid) and ranitidine (e.g., Zantac). A dense web of functionalities between histamine and H2RAs, on the one hand, and downstream cellular pathways, on the other hand, links disparate physiological pathways in gastrointestinal contexts (e.g., acid reduction) to the dysregulated inflammatory cascades (cytokine storm) underlying the pathophysiology of COVID-19. Is famotidine beneficial in treating COVID-19? This question remains unresolved, though not for lack of effort: over 10 studies have examined the potential therapeutic value of famotidine in COVID-19, but have found conflicting results (pro-famotidine, anti-famotidine, and neutral). Given the contradictory reports, we have undertaken the new analysis reported herein. Notably, studies published thus far rest upon substantially smaller datasets than drawn upon in the present work. We analyzed a cohort of 22,560 COVID-19 patients taking H_1_/H_2_ receptor antagonists, focusing on 1,379 severe cases requiring respiratory support. We analyzed outcomes for treatment with the H1RAs loratadine (e.g., Claritin) and cetirizine (e.g., Zyrtec), the H2RA famotidine, aspirin, and a famotidine & aspirin combination. For cases that reached the point of respiratory support, we found a significantly reduced fatality risk for famotidine treatment. We did not detect a benefit from dual-histamine receptor blockade (concurrently targeting H_1_ and H_2_ receptors). Notably, famotidine combined with aspirin did exhibit a significant synergistic survival benefit (odds ratio of 0.55). The relative risk for death decreased by 32.5%--an immense benefit, given the more than 2.6 million COVID-19-related deaths thus far. We found lower levels of serum markers for severe disease (e.g., C-reactive protein) in famotidine users, consistent with prior findings by others and with a role for famotidine in attenuating cytokine release. The large, international, multi-center retrospective study reported here, sampling over 250,000 COVID-19 cases, hopefully helps clarify the possible value of clinically-approved histamine antagonists such as famotidine. Given these findings, alongside the cost-effectiveness and mild side-effects of popular drugs like famotidine and aspirin, we suggest that further prospective clinical trials, perhaps utilizing the aspirin combination reported here, are advisable.

## Introduction

The coronavirus disease 2019 (COVID-19) pandemic has driven great interest in the therapeutic potential of repurposed drugs with well-established gastroenterological benefits, including those that reduce acid production and secretion in the gastric mucosa.^1^ Acid-reducing drugs belong to two main classes, based on their mechanisms of action (MOA): (i) Proton-pump inhibitors (PPIs) sterically block H^+^/K^+^-ATPase pumps, impeding the final step of acid release. (ii) Histamine H_2_ receptor antagonists (H2RA) competitively bind the H2R, a type of G-protein coupled receptor (GPCR),^2^ and block its natural stimulation by histamine; famotidine (e.g., Pepcid^®^) and ranitidine (e.g., Zantac^®^) are exemplary H2RAs.

An intricate and dense web of functional linkages exists between histamine and H2RAs, on the one hand, and disparate physiological pathways on the other hand. These pathways include gastrointestinal contexts (acid reduction) as well as the dysregulated inflammatory cascades (cytokine storm) that likely underlie much of the pathophysiology of COVID-19; these linkages have been reviewed recently. ^2,3^ Here, we simply note that mounting cellular-scale evidence suggests that the mechanistic basis for a putative role of famotidine in COVID-19 likely involves the roles of H2RA versus, for instance, direct binding of famotidine to the viral protease 3CL^pro^ (and resultant inhibition)^4^, as was originally suspected from molecular docking studies.

Given the many conceivable mechanistic linkages alluded to above, is famotidine beneficial in treating COVID-19, as gauged by outcomes involving either (i) *infection transmissibility*, (ii) *disease severity indicators* (e.g., likelihood of cases reaching the point of ventilation, WHO severity index), or (iii) *mortality rates*? Unfortunately this question remains unresolved, though not for lack of effort: since a pioneering report^5^ of positive clinical outcomes with famotidine use in COVID-19, the past year has seen over 10 studies interrogate the potential therapeutic benefits of famotidine. Many reports concluded in favor of famotidine use (e.g., refs ^5-9^), others found little to no association between famotidine (or PPIs) and measures such as 30-day mortality^10-12^ (ranitidine, another H2RA, was scrutinized in one study^13^), and a recent study found a negative association for both PPIs and famotidine.^14^ Each of these independent studies were retrospective and observational; most were cohort-based, with some as case-series (e.g., symptom tracking across longitudinal data); most evaluated inpatient cases; and most attempted to account for confounders and other biases (e.g., via propensity-score matching). Given the conflicting reports thus far, particularly the evidence that suggests a beneficial impact of famotidine on mortality and overall disease progression (e.g., mechanical ventilation), we have undertaken a new analysis, reported herein.

We note that all three parallel tracks of findings—those indicating for and against famotidine, as well as neutral (i.e., no association)—rest upon substantially smaller datasets than were drawn upon in our present work. Are any beneficial effects of famotidine detectable on a population-wide, international scale? Is it synergistic to treat with famotidine in conjunction with aspirin, a general-purpose anti-inflammatory? Does famotidine use correlate with any measurable parameters that may serve as biomarkers, perhaps offering mechanistic clues (e.g., serum C-reactive protein [CRP] levels as a proxy for inflammation and the cytokine storm)? This work seeks to address these questions.

## Methods

We retrieved data from the COVID-19 Research Network supplied by TriNetX, comprising ≈400M patients from 130 health care organizations in 30 countries. TriNetX provides a global federated health research network providing access to electronic medical records (diagnoses, procedures, medications, laboratory values, genomic information), and the TriNetX platform uses only aggregated counts and statistical summaries of de-identified information; no protected health information (PHI) or personal data is made available on the platform. This work was reviewed by our IRB board (UVA IRB tracking ID #23100), who determined that this project did not meet the criteria for Human Subject Research; therefore, no additional IRB submission/review was deemed necessary (by the IRB) to proceed with this project. We analyzed a cohort of 22,560 COVID-19 patients taking H_1_/H_2_receptor antagonists, with a special focus on 1,379 severe cases requiring respiratory support (see CONSORT flow diagram, Supplemental Figure 1). We defined death as the primary outcome, and, in order to try to mitigate confounder bias, we performed propensity score matching to achieve stratified and balanced sub-cohorts across age and gender; specifically, we balanced cohorts using a nearest-neighbor greedy matching algorithm with a caliper of 0.25 times the standard deviation. Measures of association, risk ratios (RRs) and odds ratios (ORs), along with their respective 95% CIs, were calculated. Kaplan-Meier survival curves were also computed for each analysis.

**Figure 1.**
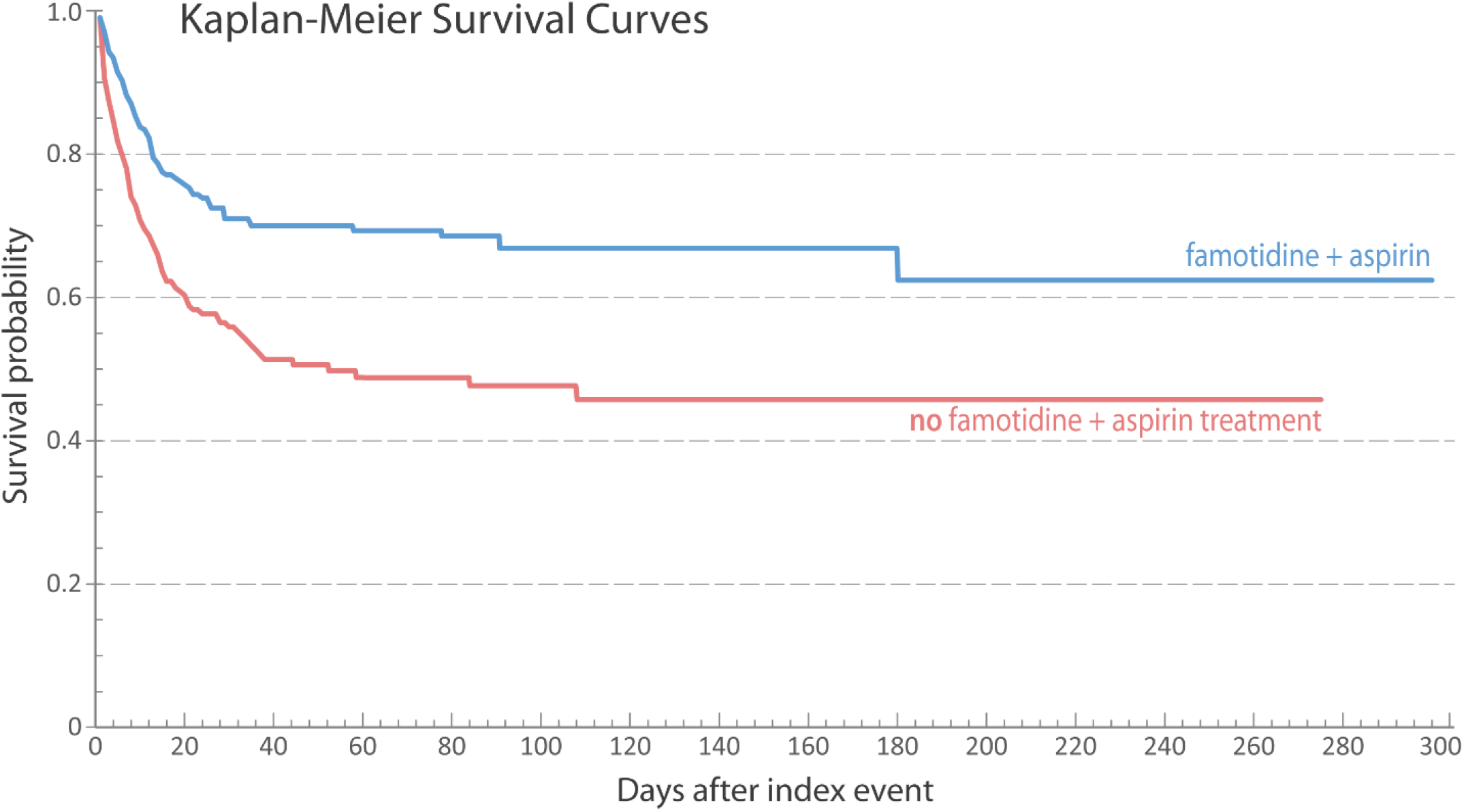
**Kaplan-Meier survival curves** are shown for COVID-19 patients with (blue) or without (red) the dual combination treatment of famotidine and aspirin.

## Results & Discussion

We statistically analyzed outcomes for treatment with (i) the H1R antagonists loratadine (e.g., Claritin^®^) and cetirizine (e.g., Zyrtec^®^), (ii) the H2RA famotidine, (iii) the general-purpose medication aspirin, and (iv) a combination of famotidine & aspirin. For cases that reached the point of respiratory support, we found a significantly reduced fatality risk for famotidine treatment (OR 0.73, CI 0.57 to 0.94; Table 1, Supplemental Files 1-4). Hogan et al.^6^ recently suggested that dual-histamine receptor blockade, concurrently targeting both the H_1_ and H_2_ receptors, can improve COVID-19 clinical outcomes; however, significant improvements were not found in our cohorts, versus famotidine alone (OR 0.75, CI 0.39 to 1.46; Supplemental Files 5-8). Notably, and perhaps unexpectedly, the combination of famotidine and aspirin (344 severe cases before matching) did exhibit a significant synergistic survival benefit (OR 0.55, CI 0.39 to 0.78; Supplemental Files 9-12). The risk ratio for death decreased by 32.5%—an immense benefit, given the more than 2.6 million COVID-19–related deaths thus far.

**Table 1:** **Statistical outcomes for patients requiring respiratory support**, considering use/disuse of (i) H_1_- or H_2_-receptor antagonists or aspirin, as well as (ii) a combination treatment with famotidine and aspirin.

| Drug compound<br>[H<sub>1</sub> or H<sub>2</sub> antagonist] | Number of pa-<br>tients in cohort<br>(after matching) | Outcome:<br>Death | Odds ratio<br>(OR) | Confidence interval<br>(CI 95%) | Hazard ratio<br>(HR) |
| --- | --- | --- | --- | --- | --- |
| Loratadine [H<sub>1</sub>] | 88 | 29 | 1.00 | 0.55–1.87 | 0.84 |
| Cetirizine [H<sub>1</sub>] | 95 | 25 | 0.85 | 0.45–1.61 | 0.80 |
| Famotidine [H<sub>2</sub>] | 563 | 161 | 0.73 | 0.57–0.94 | 0.75 |
| Aspirin (Asp) | 527 | 165 | 0.79 | 0.61–1.02 | 0.71 |
| Famotidine + Asp | 305 | 83 | 0.55 | 0.39–0.78 | 0.53 |

Can these findings be reconciled with the recent studies (mentioned above) of famotidine and COVID-19? Janowitz et al.^7^ reported a case-series of 10 non-hospitalized COVID-19 patients for whom self-administration of famotidine had uniformly beneficial impact on disease trajectories, based on quantitative symptom-tracking across longitudinal data. Retrospective, single-center^5,6,8^ studies also found promising results, in terms of reduced risk of clinical deterioration leading to intubation or death, for famotidine usage in 83 and 84 hospitalized COVID-19 patients, corresponding to 9.5 and 5.1% of the analyzed cohorts of patients, respectively.^5,8^ Notably, both Freed-berg et al.^5^ and Mather et al.^8^ found lower levels of serum markers for severe disease (e.g., ferritin, C-reactive protein, or procalcitonin) in the famotidine group, consistent with our findings and with a potential role for this H2RA in attenuating cytokine release. Finally, a new systematic review and analysis (of published reports) suggests that famotidine may be beneficial^15^, while two other recent meta-analyses are either neutral or (statistically) inconclusive^16,17^ as regards famotidine use in COVID-19.

If indeed famotidine is of therapeutic benefit in (most) COVID-19 cases, what might be its etiological basis? Beyond what is alluded to above, the molecular-scale MOAs that can be envisioned are covered in other recent work.^18^ Here, we simply reiterate that famotidine or other H2RAs may aid COVID-19 patients because of the capacity of such compounds to attenuate pro-inflammatory pathways that become dysregulated upon an infection (cytokine storms activate pro-fibrotic pathways; lung damage eventually results). Thus, a role for famotidine in COVID-19 may stem from cellular mechanisms that are rather indirect and unrelated to this compound’s classic therapeutic role in gastroenterology.

As SARS-CoV-2 infection rates continue surging worldwide, we desperately need more data on potential therapies. The large, international, multi-center retrospective study reported here, sampling over 250,000 COVID-19 cases, hopefully helps clarify the potential benefit of clinically-approved histamine antagonists such as famotidine. We anticipate that at least three prospective, randomized, controlled clinical trials currently underway—NCT04504240, NCT04370262 and NCT-04545008—will illuminate famotidine’s potential therapeutic profile. Given the findings reported here, alongside the cost-effectiveness and mild/manageable side-effects of OTC drugs like famotidine and aspirin, we suggest that further prospective clinical trials—perhaps utilizing the aspirin combination reported here—are advisable.

## Data Availability

Data can be made available to researchers upon request.

## Document information

## Abbreviations

CI,: confidence interval;
CONSORT,: Consolidated Standards of Reporting Trials;
COVID-19,: coronavirus disease 2019;
OR,: odds ratio;
PPI,: proton-pump inhibitor;
RR,: risk ratio;
SARS-CoV-2,: severe acute respiratory syndrome coronavirus 2;

## Additional notes

The main text includes 1 figure, 1 table, and is accompanied by several items of Supplementary Material.

## Acknowledgements

We thank Dr Flaminia Coluzzi (Sapienza University of Rome, Italy) for contributions to the project and feedback on the manuscript.

## Patient consent for publication

Not required; see Methods section for IRB review.

## Supplementary Materials

The following supplementary figures, tables, and data files accompany this manuscript:

- Supplemental Figure 1: CONSORT Flow Diagram
- Supplemental Table 1:Lab Values and Standard Deviations for Serum Levels of C-reactive Protein
- Supplemental File 1: Measures of Association Data Graph for Famotidine
- Supplemental File 2: Measures of Association Data Table for Famotidine
- Supplemental File 3: Kaplan-Meier Raw Data Graph for Famotidine
- Supplemental File 4: Kaplan-Meier Raw Data Table for Famotidine
- Supplemental File 5: Measures of Association Data Graph for H1 and H2
- Supplemental File 6: Measures of Association Data Table for H1 and H2
- Supplemental File 7: Kaplan-Meier Raw Data Graph for H1 and H2
- Supplemental File 8: Kaplan-Meier Raw Data Table for H1 and H2
- Supplemental File 9: Measures of Association Data Graph for Famotidine and Asp
- Supplemental File 10: Measures of Association Data Table for Famotidine and Asp
- Supplemental File 11: Kaplan-Meier Raw Data Graph for Famotidine and Asp
- Supplemental File 12: Kaplan-Meier Raw Data Table for Famotidine and Asp

---

**Supplemental Figure 1:**
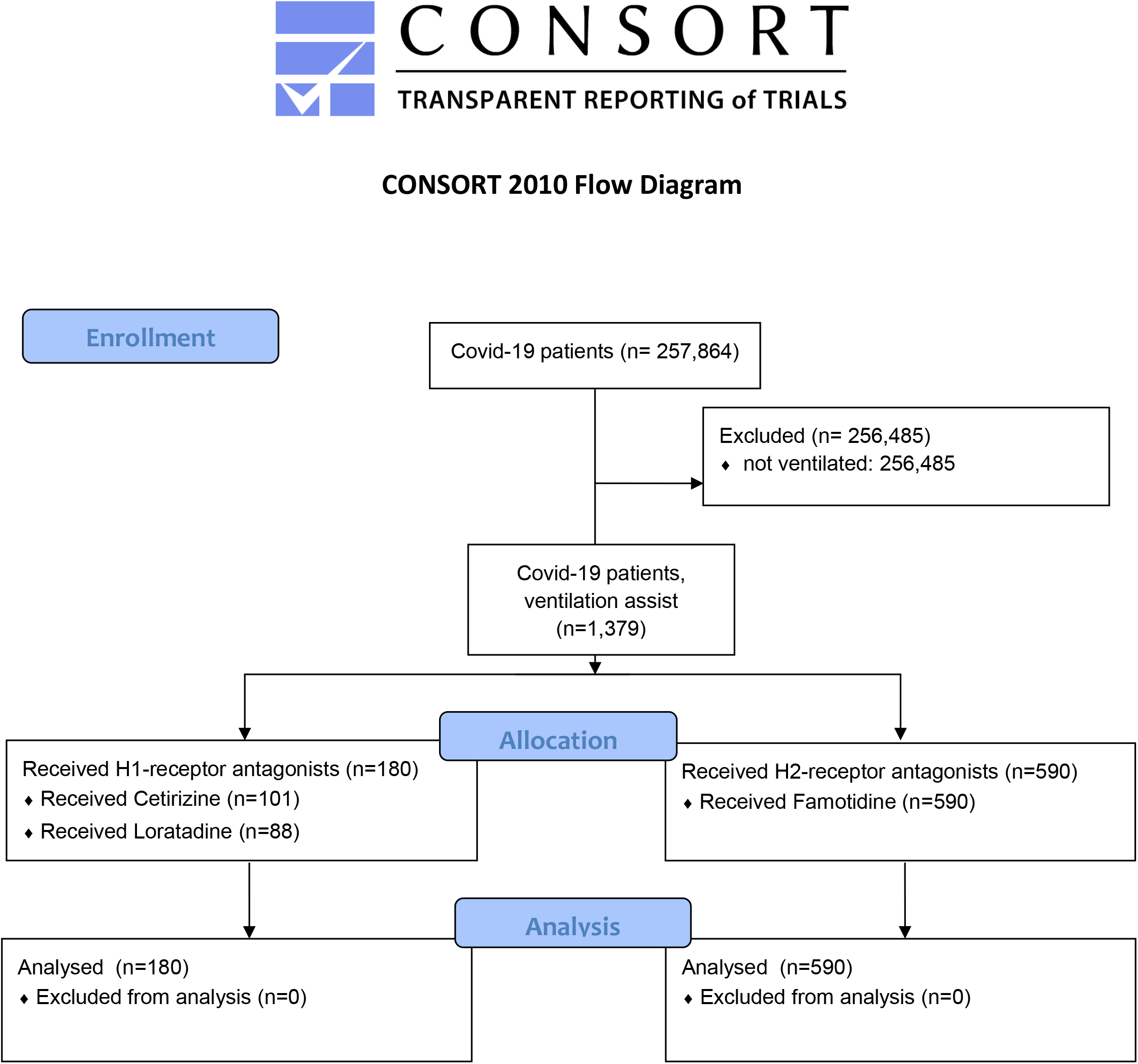
CONSORT flow diagram for this study.

**Supplemental Table 1:** Lab values and standard deviations (SD) for serum levels of C-reactive protein (CRP) are given for COVID-19 patients in this study; note that serum CRP levels exceeding ≈10 mg/l are generally indicative of severe infection or disease. Here, we consider six cohorts: Comparing patients who either (1a) died or (1b) survived shows large differences in mean CRP values, as does a comparison of (2a) ventilated versus (2b) non-ventilated patients; perhaps unsurprisingly, those requiring ventilation had CRP levels nearly 2-fold higher than the non-ventilated sub-cohort. Among ventilated patients, the combination of aspirin and famotidine (cohort 3b) is associated with less of a decrease in CRP values (below cohort 3a), versus the differences within other cohort pairs (i.e., cohorts 1a/b and 2a/b); this trend is actually stronger than suggested here, as this cohort is six years older on average and has more comorbidities (data not shown).

| Cohort | Inclusion criteria | CRP (mg/l) | SD | Sub-cohort difference |
| --- | --- | --- | --- | --- |
| COVID-19 1a | deceased | 100.0 | 107.0 | 65.7 |
| 1b | survived | 34.3 | 58.0 |  |
| COVID-19 2a | ✓ ventilation | 74.9 | 98.5 | 35.3 |
| 2b | ✗ ventilation | 39.6 | 65.6 |  |
| COVID-19 3a | ventilation ✗ aspirin, ✗ famotidine | 81.7 | 104.0 | 13.3 |
| 3b | ventilation ✓ aspirin, ✓ famotidine | 68.4 | 95.4 |  |

