## Supplementary material for "Clinical Evidence for Improved Outcomes with Histamine Antagonists and Aspirin in 22,560 COVID-19 Patients": Supp Files 1-4: _Read_Me.txt

CONTENTS OF THIS FILE
---------------------

 * Link to Results
 * Study Metadata
 * List of Files Included in Export
 * Cohort 1 Inclusion/Exclusion Criteria
 * Cohort 2 Inclusion/Exclusion Criteria

LINK TO RESULTS
---------------

 * https://live.trinetx.com/tnx/study/68098/analytics/5fa115b5802aa409b6298787/outcomes/results


STUDY METADATA
--------------

 * Study Name: repurposing
 * Analysis Name: +/- famotidine (vent)
 * Analysis Type: Compare Outcomes
 * Cohort 1: CovRedef+famotidine+vent
 * Cohort 2: CovRedef-famotidine+vent
 * Run by: Robert Preissner
 * Run on: Nov 13, 2020, 9:39 am
 * Network Name: COVID-19 Research Network
 * Patients on Network at Time of Analysis: 70,948,314
 * HCOs on Network at Time of Analysis: 55
 * HCOs Online for Analysis: 54
 * Last Data Refresh: Nov 13, 2020, 6:14 am
 * Outcome 1 Name: death


LIST OF FILES INCLUDED IN EXPORT
--------------------------------

 * Outcome_1_Result_b_KM_table.png
 * Outcome_1_Result_a_MOA_table.png
 * Baseline_Patient_Characteristics.png
 * Baseline_Patient_Characteristics.csv
 * Propensity_Score_Density_Graph_Small.png
 * Outcome_1_Result_b_KM_graph_small.png
 * Outcome_1_Result_a_MOA_graph_small.png
 * Propensity_Score_Density_Graph_Large.png
 * Propensity_Score_Density_Graph.csv
 * Outcome_1_Result_b_KM_graph_large.png
 * Outcome_1_Result_b_KM_table.csv
 * Outcome_1_Result_b_KM_graph.csv
 * Outcome_1_Result_a_MOA_graph_large.png
 * Outcome_1_Result_a_MOA_table.csv
 * Outcome_1_Result_a_MOA_graph.csv


COHORT 1 INCLUSION/EXCLUSION CRITERIA
-------------------------------------

 * Event 1A This event occurred since 11 months ago

 * Must Have: 1014859 Ventilation assist and management, initiation of pressure or volume preset ventilators for assisted or controlled breathing AND 4278 famotidine AND (B97.29 Other coronavirus as the cause of diseases classified elsewhere OR 9088 SARS coronavirus 2 and related RNA [Presence] [Positive, ever] OR 94534-5 SARS-CoV-2 (COVID19) RdRp gene [Presence] in Respiratory specimen by NAA with probe detection [Positive, ever] OR 94500-6 SARS-CoV-2 (COVID19) RNA [Presence] in Respiratory specimen by NAA with probe detection [Positive, ever] OR 94309-2 SARS-CoV-2 (COVID19) RNA [Presence] in Unspecified specimen by NAA with probe detection [Positive, ever] OR 94533-7 SARS-CoV-2 (COVID19) N gene [Presence] in Respiratory specimen by NAA with probe detection [Positive, ever] OR 94315-9 SARS-CoV-2 (COVID19) E gene [Presence] in Unspecified specimen by NAA with probe detection [Positive, ever] OR 9089 SARS coronavirus 2 IgG IgM Ab [Presence] in Serum or Plasma [Positive, ever] OR 94563-4 SARS coronavirus 2 IgG Ab [Presence] in Serum or Plasma by Immunoassay [Positive, ever] OR 94316-7 SARS-CoV-2 (COVID19) N gene [Presence] in Unspecified specimen by NAA with probe detection [Positive, ever] OR 41458-1 SARS coronavirus RNA [Presence] in Unspecified specimen by NAA with probe detection [Positive, ever] OR 94507-1 SARS coronavirus 2 IgG Ab [Presence] in Serum or Plasma by Rapid immunoassay [Positive, ever] OR 94762-2 SARS coronavirus 2 Ab [Presence] in Serum or Plasma by Immunoassay [Positive, ever] OR 94505-5 SARS-CoV-2 (COVID19) IgG Ab [Units/volume] in Serum or Plasma by Immunoassay [≥ 1 units, most recent value] OR 94564-2 SARS coronavirus 2 IgM Ab [Presence] in Serum or Plasma by Immunoassay [Positive, ever] OR 94559-2 SARS coronavirus 2 ORF1ab region [Presence] in Respiratory specimen by NAA with probe detection [Positive, ever] OR 94547-7 SARS-CoV-2 (COVID19) IgG+IgM Ab [Presence] in Serum or Plasma by Immunoassay [Positive, ever] OR 94531-1 SARS-CoV-2 (COVID19) RNA panel - Respiratory specimen by NAA with probe detection [≥ 0.6 units, most recent value] OR 94508-9 SARS coronavirus 2 IgM Ab [Presence] in Serum or Plasma by Rapid immunoassay [Positive, ever] OR U07.2 COVID-19, virus not identified (WHO) OR U07.1 COVID-19 OR J12.81 Pneumonia due to SARS-associated coronavirus OR B34.2 Coronavirus infection, unspecified)

 * Cannot Have: 079.89 Other specified viral infection


COHORT 2 INCLUSION/EXCLUSION CRITERIA
-------------------------------------

 * Event 1A This event occurred since 11 months ago

 * Must Have: 1014859 Ventilation assist and management, initiation of pressure or volume preset ventilators for assisted or controlled breathing AND (B97.29 Other coronavirus as the cause of diseases classified elsewhere OR 9088 SARS coronavirus 2 and related RNA [Presence] [Positive, ever] OR 94534-5 SARS-CoV-2 (COVID19) RdRp gene [Presence] in Respiratory specimen by NAA with probe detection [Positive, ever] OR 94500-6 SARS-CoV-2 (COVID19) RNA [Presence] in Respiratory specimen by NAA with probe detection [Positive, ever] OR 94309-2 SARS-CoV-2 (COVID19) RNA [Presence] in Unspecified specimen by NAA with probe detection [Positive, ever] OR 94533-7 SARS-CoV-2 (COVID19) N gene [Presence] in Respiratory specimen by NAA with probe detection [Positive, ever] OR 94315-9 SARS-CoV-2 (COVID19) E gene [Presence] in Unspecified specimen by NAA with probe detection [Positive, ever] OR 9089 SARS coronavirus 2 IgG IgM Ab [Presence] in Serum or Plasma [Positive, ever] OR 94563-4 SARS coronavirus 2 IgG Ab [Presence] in Serum or Plasma by Immunoassay [Positive, ever] OR 94316-7 SARS-CoV-2 (COVID19) N gene [Presence] in Unspecified specimen by NAA with probe detection [Positive, ever] OR 41458-1 SARS coronavirus RNA [Presence] in Unspecified specimen by NAA with probe detection [Positive, ever] OR 94507-1 SARS coronavirus 2 IgG Ab [Presence] in Serum or Plasma by Rapid immunoassay [Positive, ever] OR 94762-2 SARS coronavirus 2 Ab [Presence] in Serum or Plasma by Immunoassay [Positive, ever] OR 94505-5 SARS-CoV-2 (COVID19) IgG Ab [Units/volume] in Serum or Plasma by Immunoassay [≥ 1 units, most recent value] OR 94564-2 SARS coronavirus 2 IgM Ab [Presence] in Serum or Plasma by Immunoassay [Positive, ever] OR 94559-2 SARS coronavirus 2 ORF1ab region [Presence] in Respiratory specimen by NAA with probe detection [Positive, ever] OR 94547-7 SARS-CoV-2 (COVID19) IgG+IgM Ab [Presence] in Serum or Plasma by Immunoassay [Positive, ever] OR 94531-1 SARS-CoV-2 (COVID19) RNA panel - Respiratory specimen by NAA with probe detection [≥ 0.6 units, most recent value] OR 94508-9 SARS coronavirus 2 IgM Ab [Presence] in Serum or Plasma by Rapid immunoassay [Positive, ever] OR U07.2 COVID-19, virus not identified (WHO) OR U07.1 COVID-19 OR J12.81 Pneumonia due to SARS-associated coronavirus OR B34.2 Coronavirus infection, unspecified)

 * Cannot Have: 079.89 Other specified viral infection OR 4278 famotidine


Generated by TriNetX
