## Supplementary figures and images for "Clinical Evidence for Improved Outcomes with Histamine Antagonists and Aspirin in 22,560 COVID-19 Patients"

### Baseline_Patient_Characteristics.png

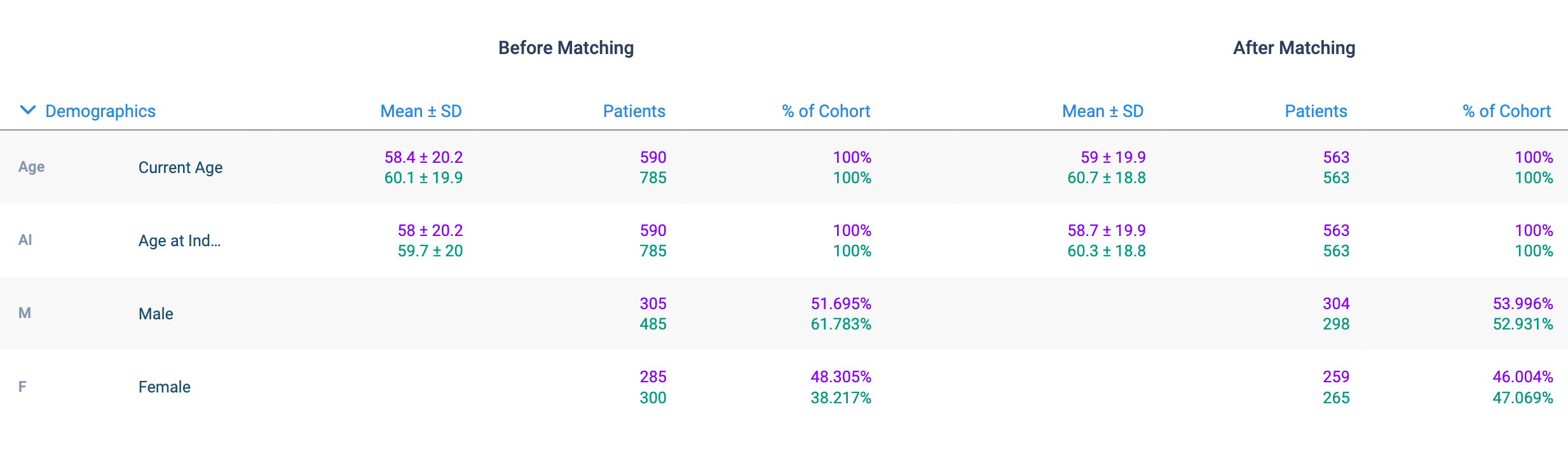

### Baseline_Patient_Characteristics.png

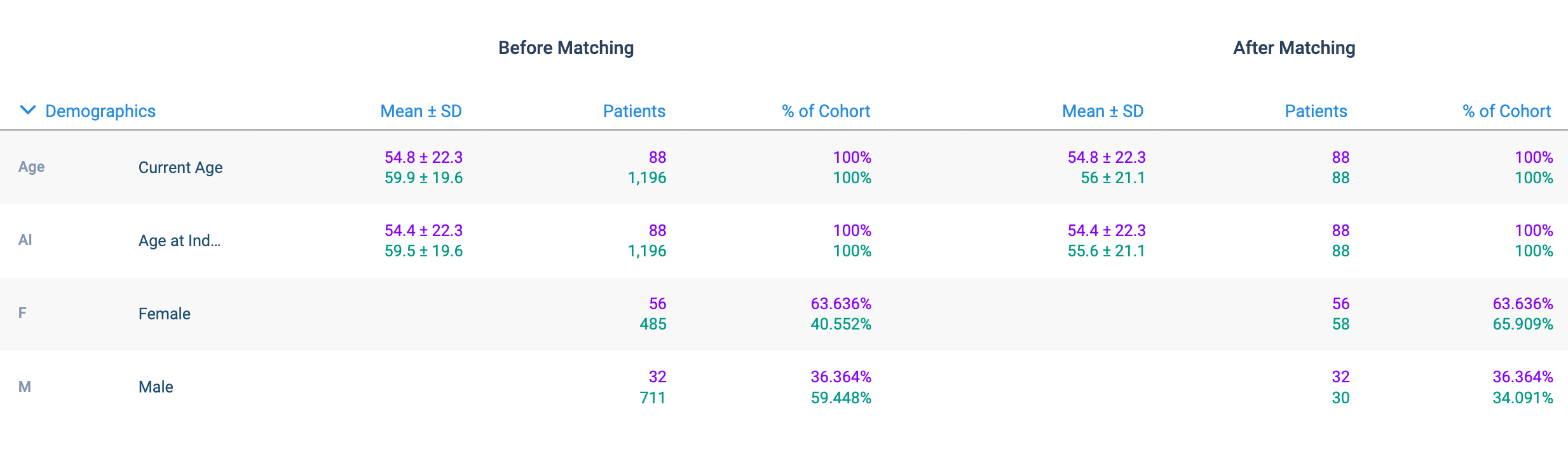

### Baseline_Patient_Characteristics.png

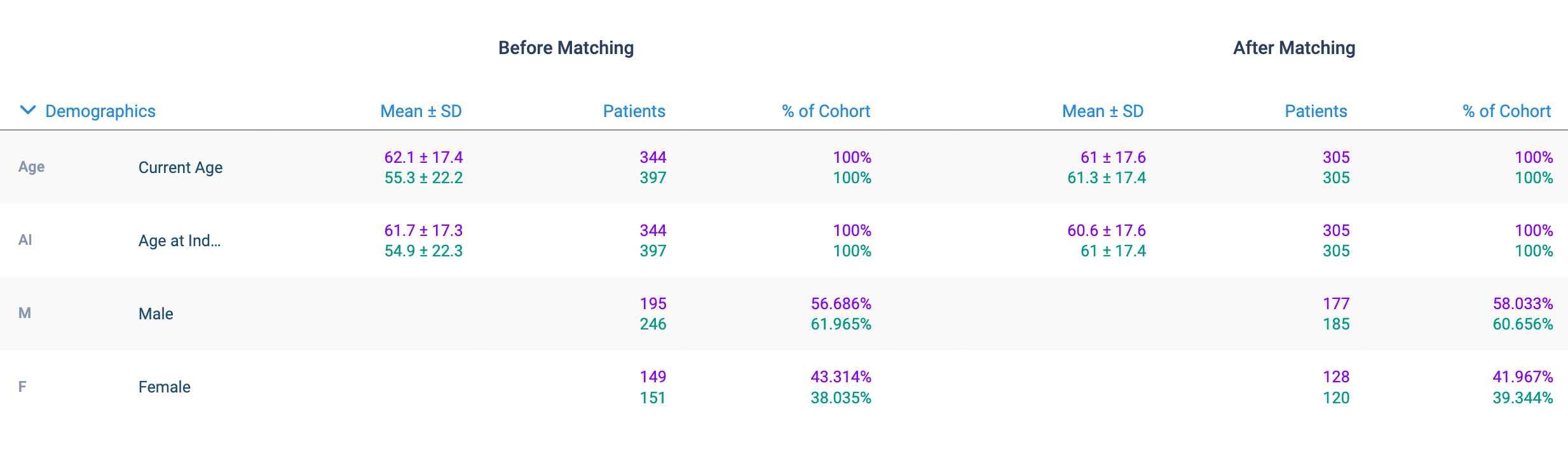

### Outcome_1_Result_a_MOA_graph_large.png

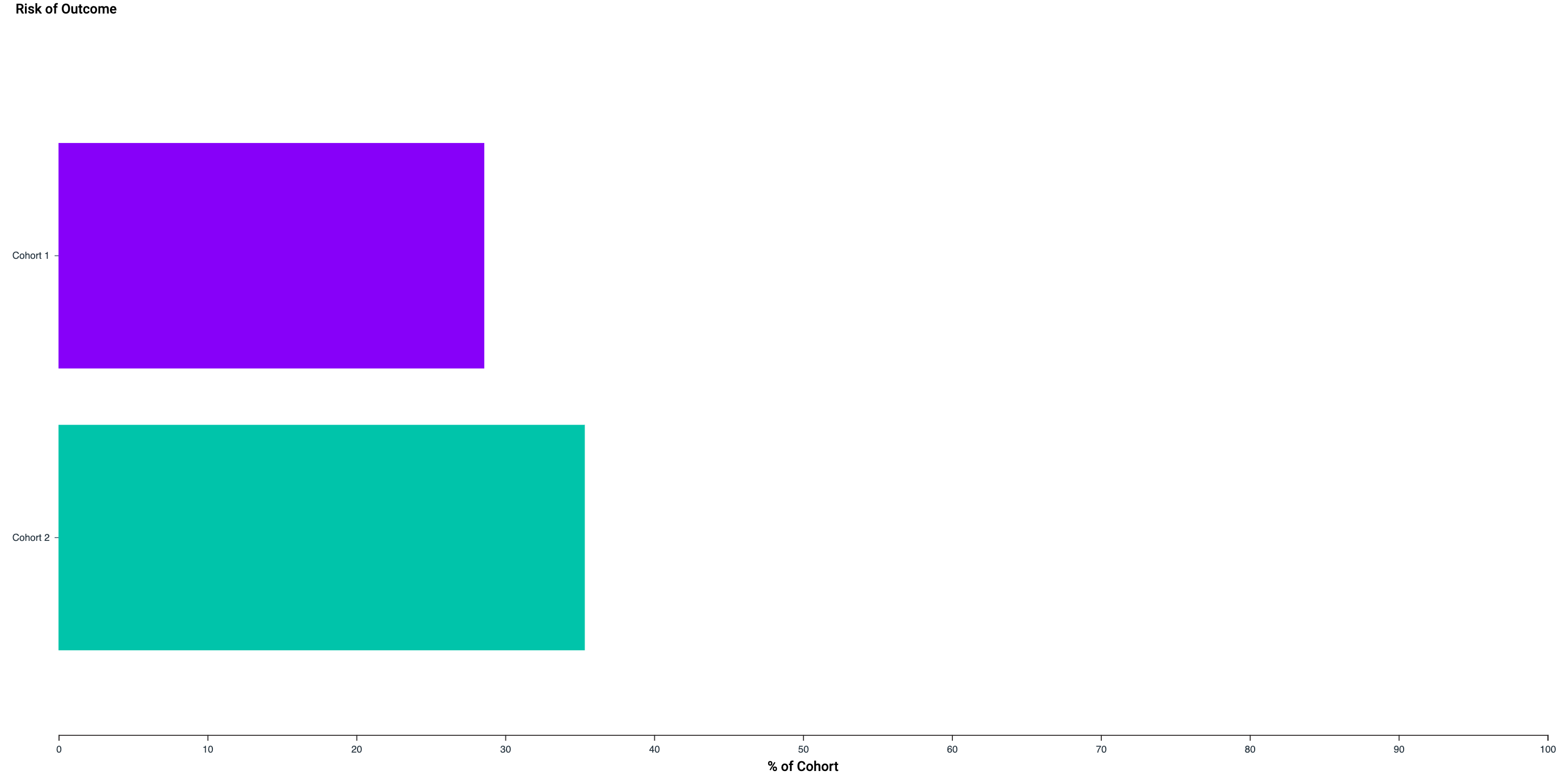

### Outcome_1_Result_a_MOA_graph_large.png

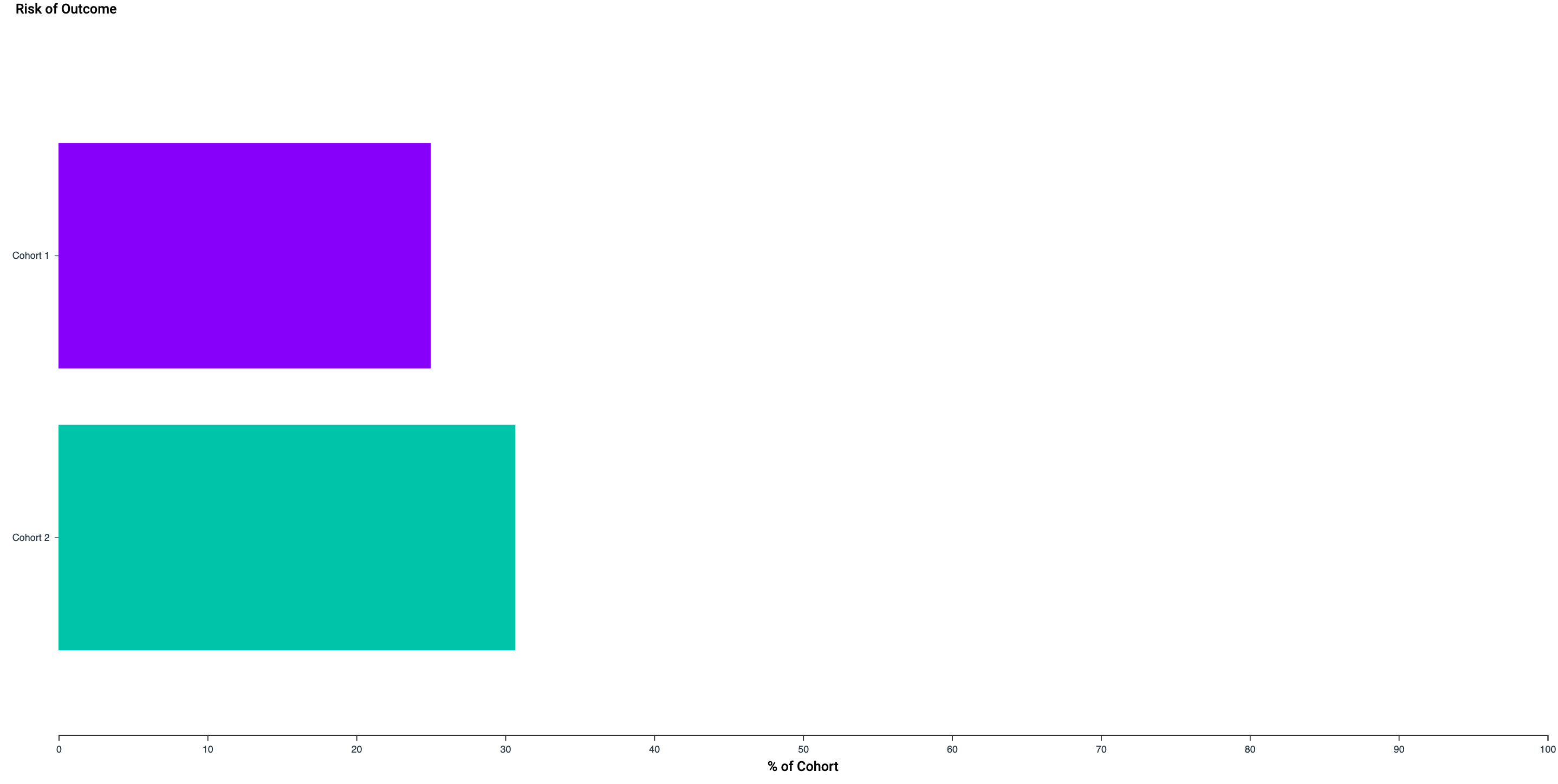

### Outcome_1_Result_a_MOA_graph_large.png

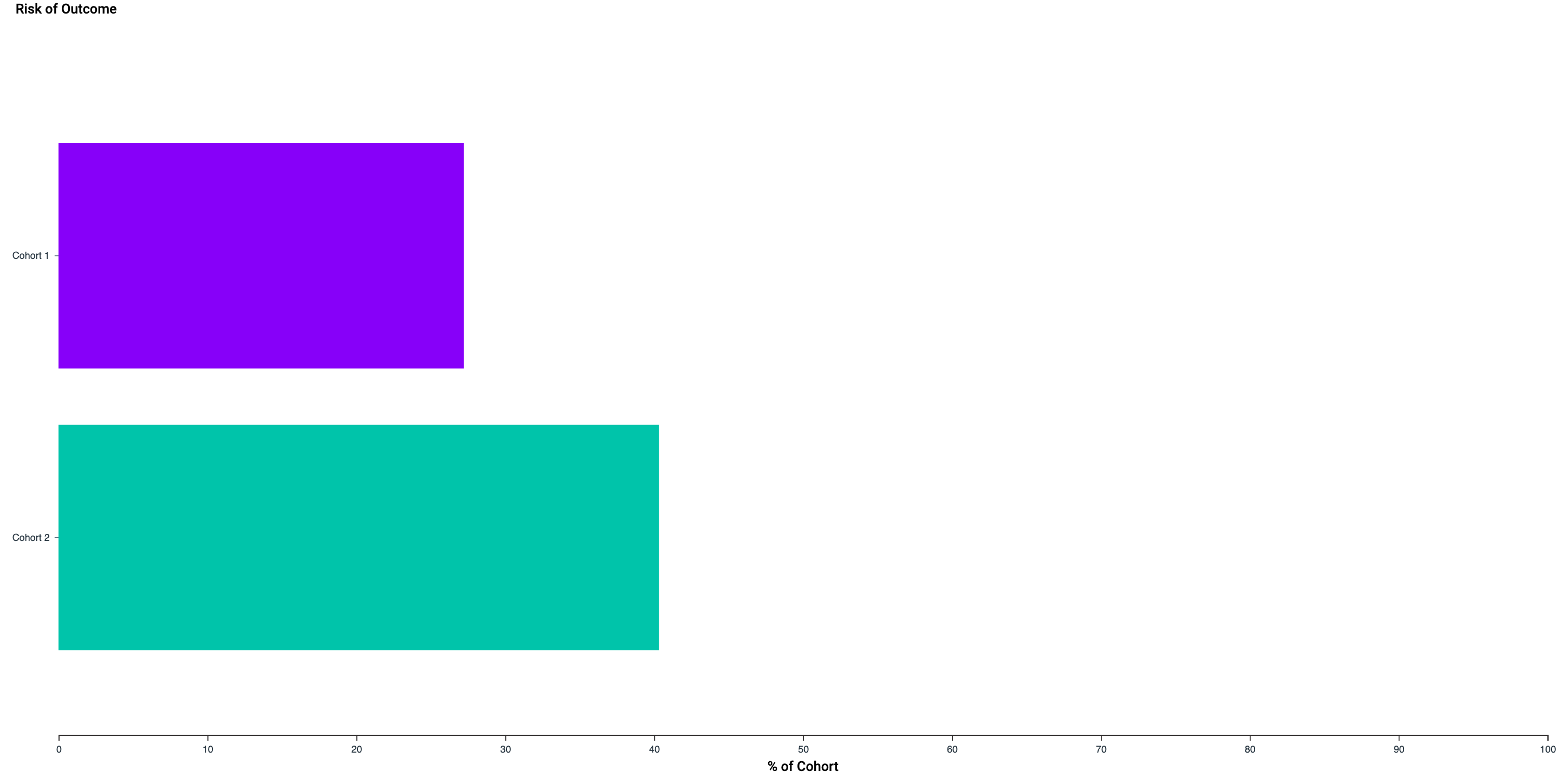

### Outcome_1_Result_a_MOA_graph_small.png

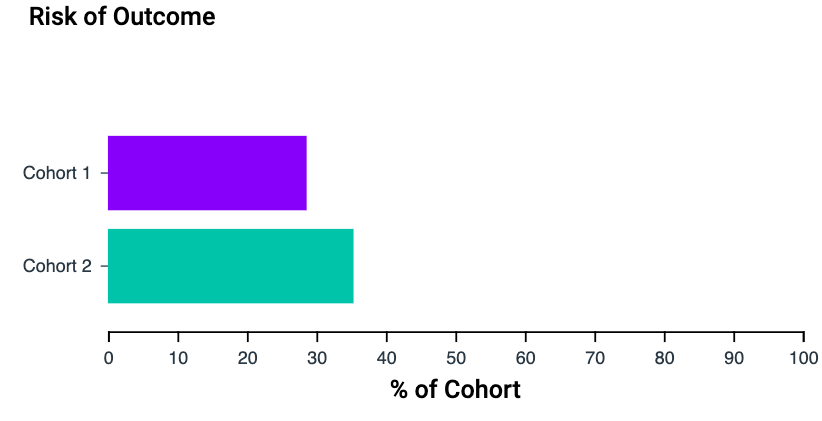

### Outcome_1_Result_a_MOA_graph_small.png

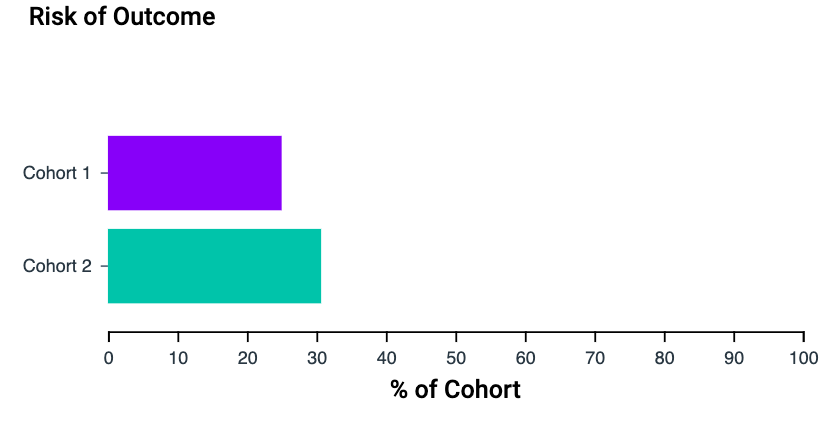

### Outcome_1_Result_a_MOA_graph_small.png

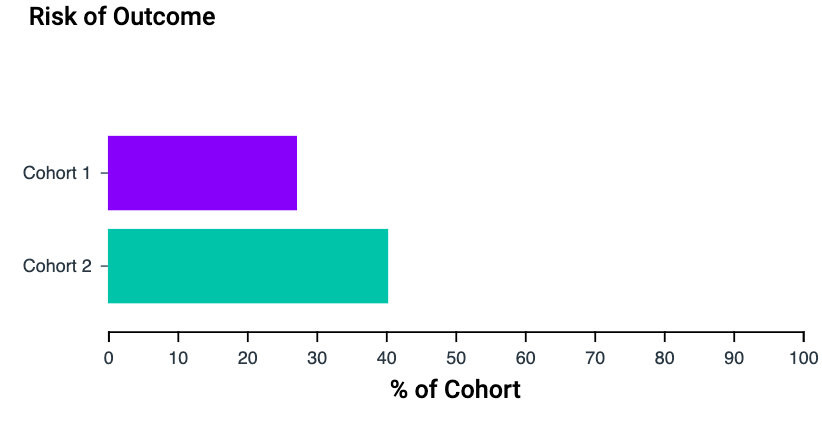

### Outcome_1_Result_a_MOA_table.png

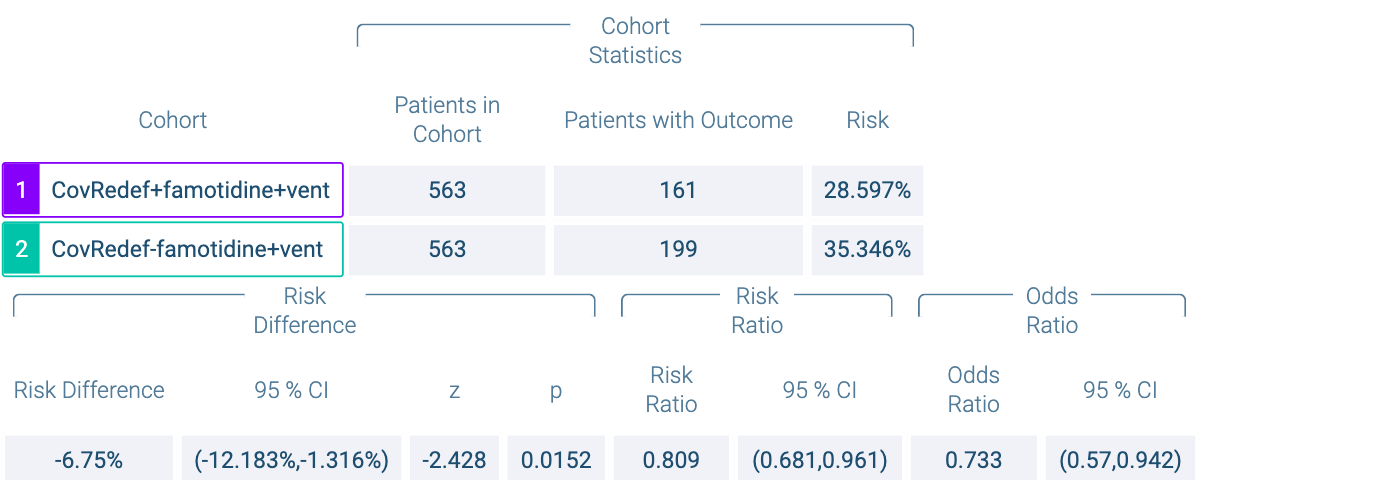

### Outcome_1_Result_a_MOA_table.png

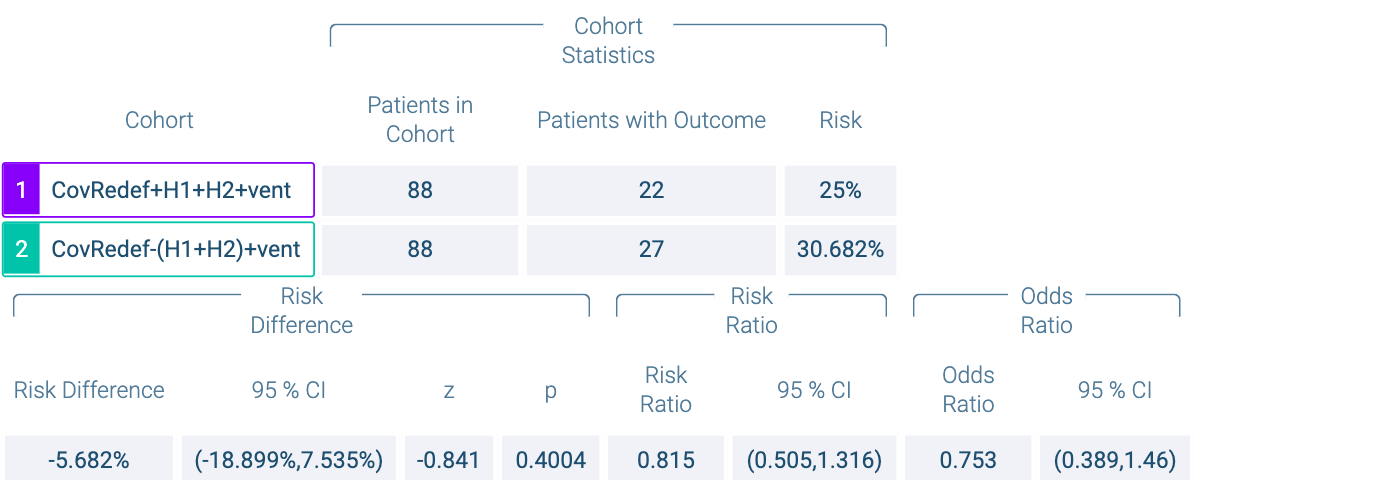

### Outcome_1_Result_a_MOA_table.png

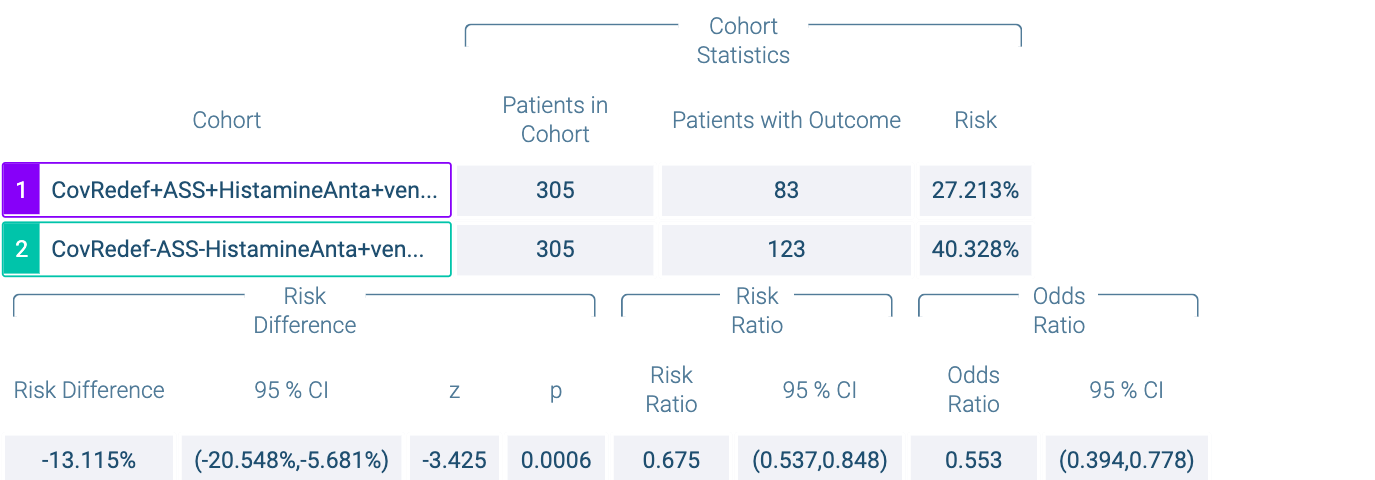

### Outcome_1_Result_b_KM_graph_large.png

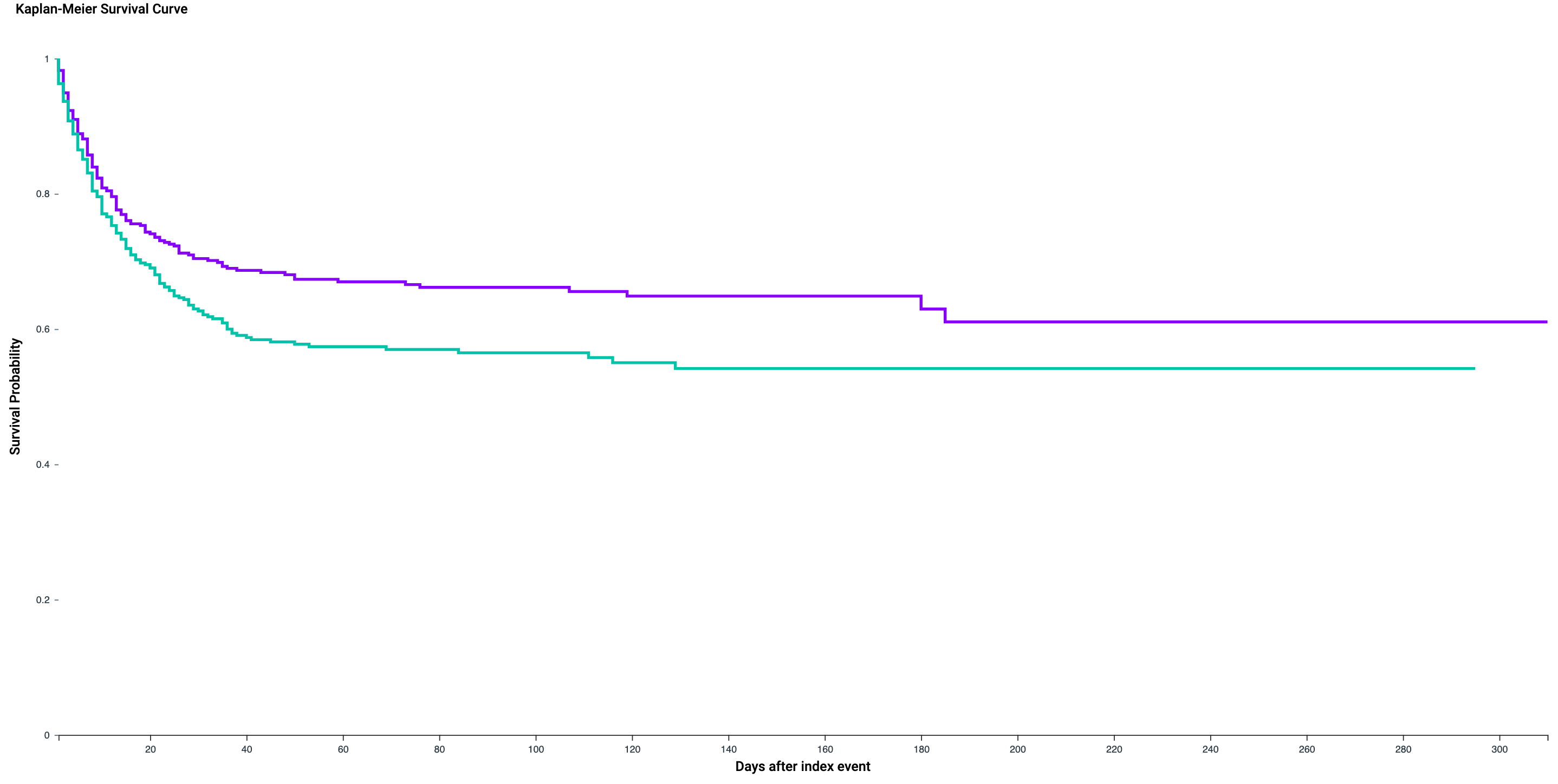

### Outcome_1_Result_b_KM_graph_large.png

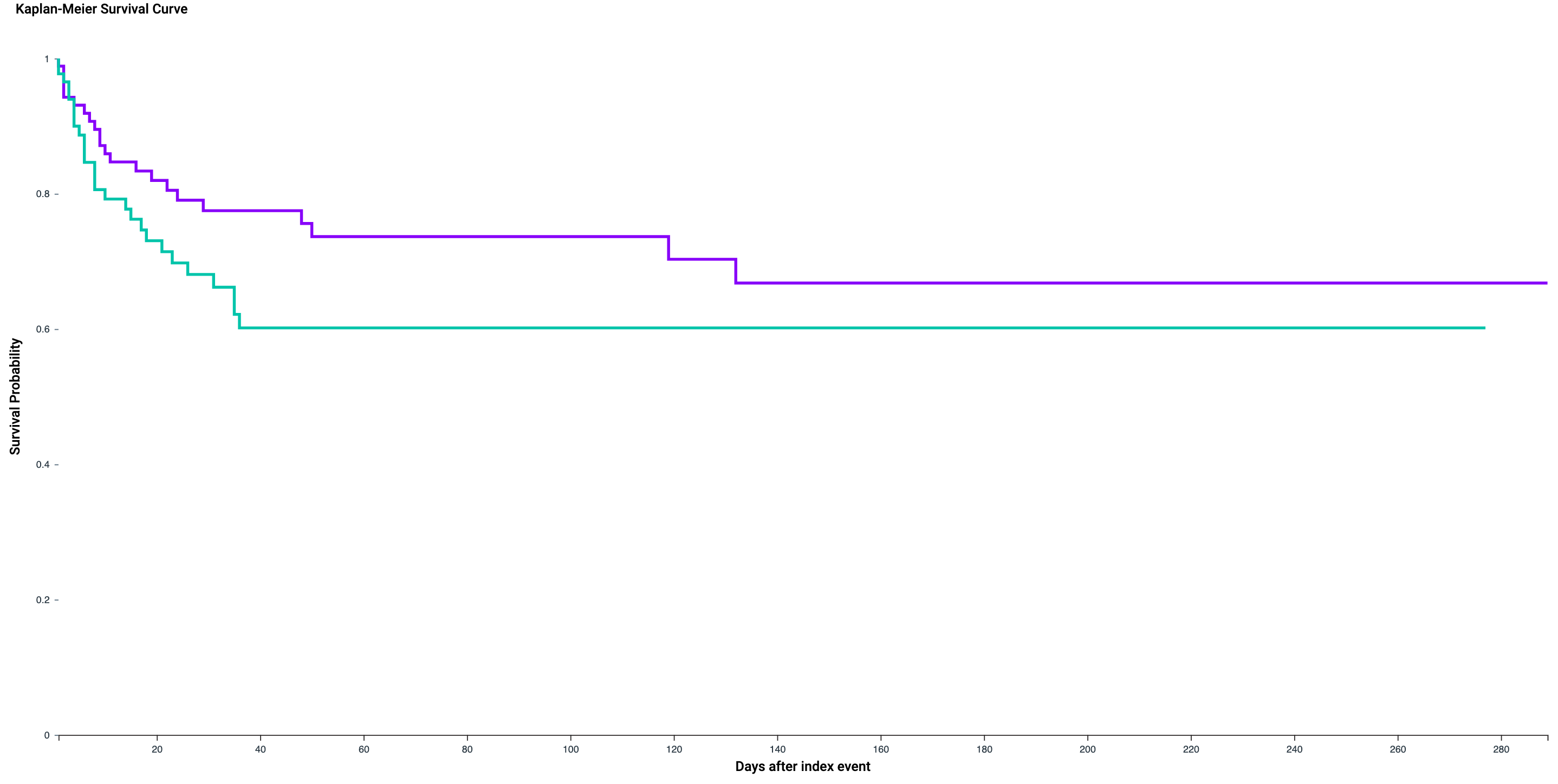

### Outcome_1_Result_b_KM_graph_large.png

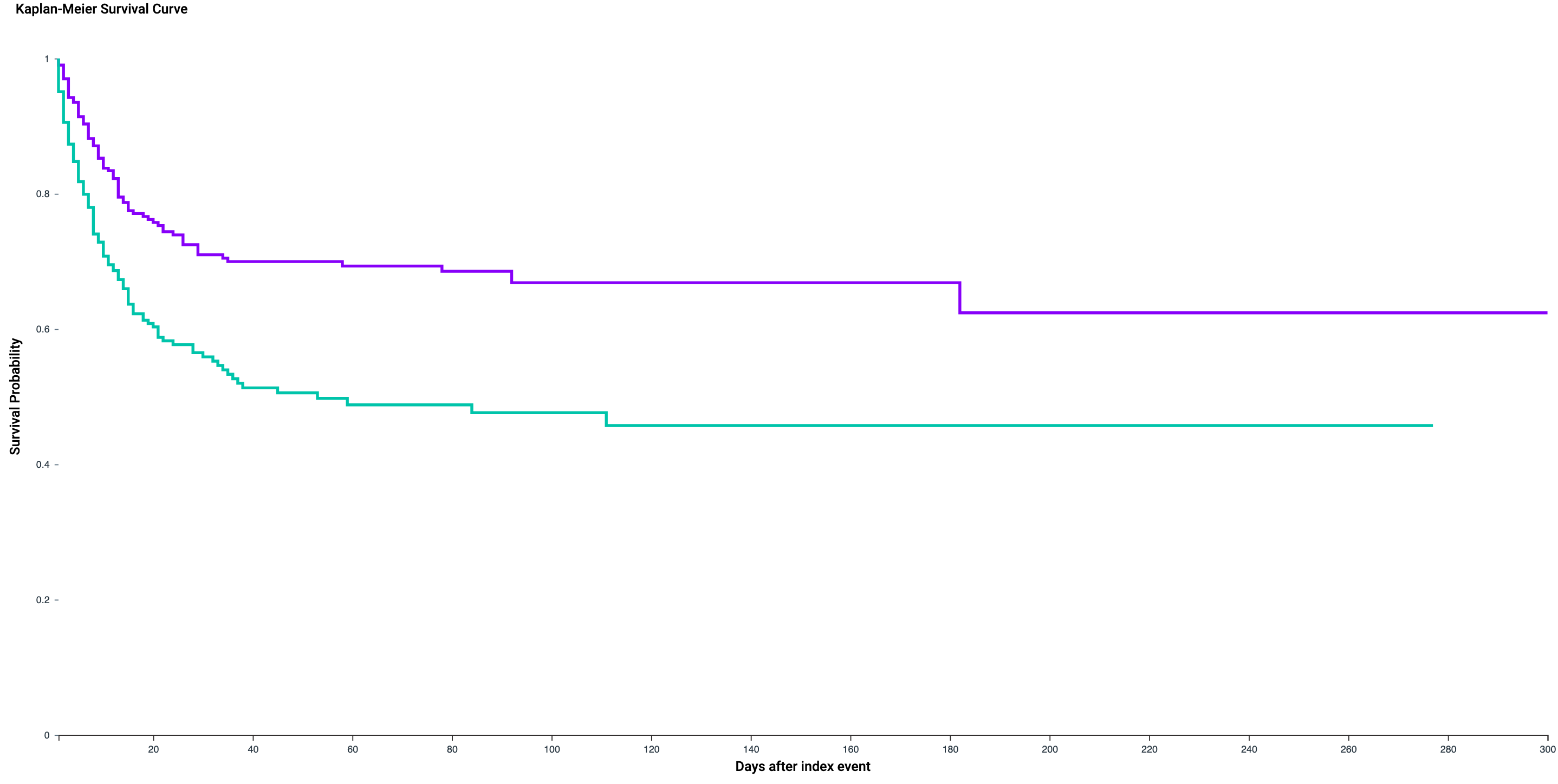

### Outcome_1_Result_b_KM_graph_small.png

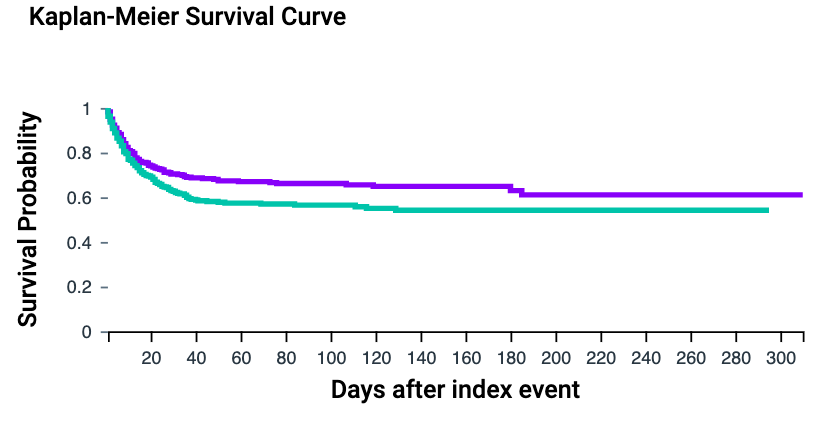

### Outcome_1_Result_b_KM_graph_small.png

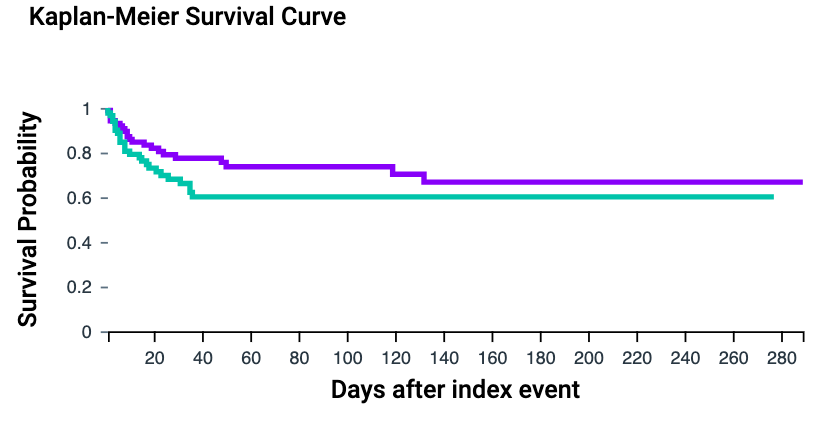

### Outcome_1_Result_b_KM_graph_small.png

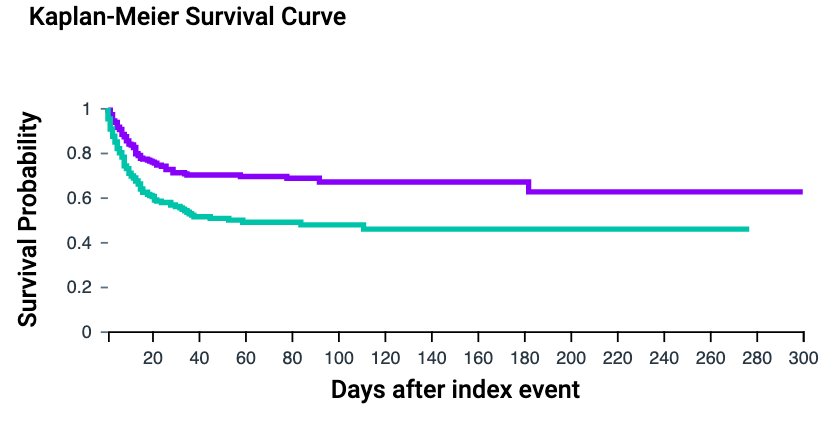

### Outcome_1_Result_b_KM_table.png

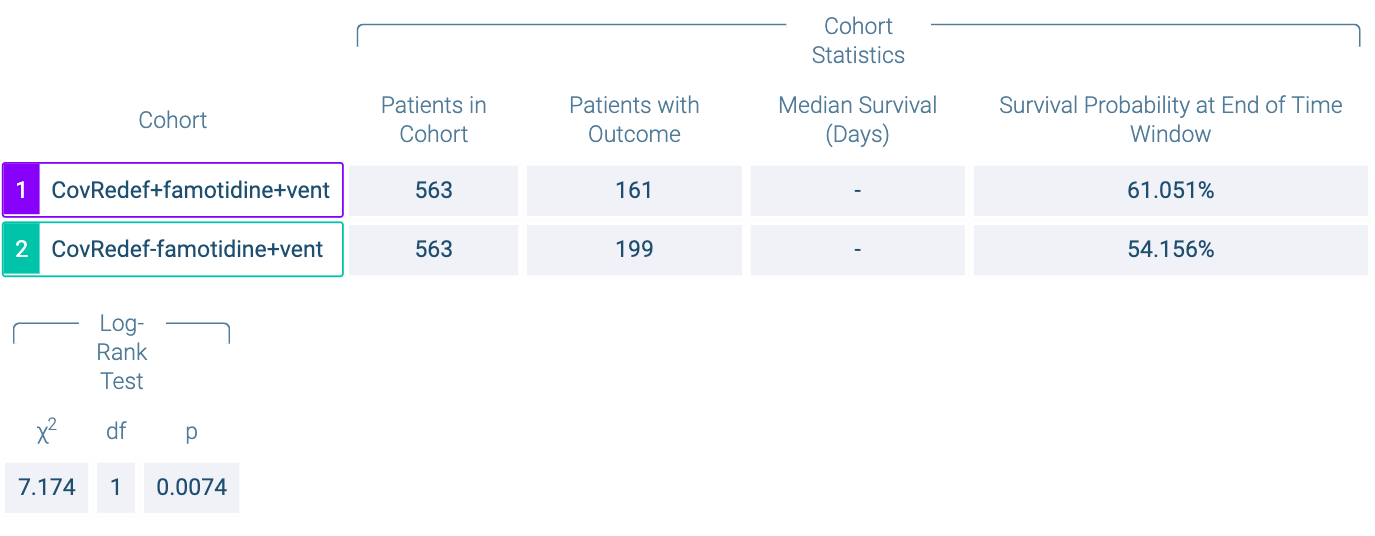

### Outcome_1_Result_b_KM_table.png

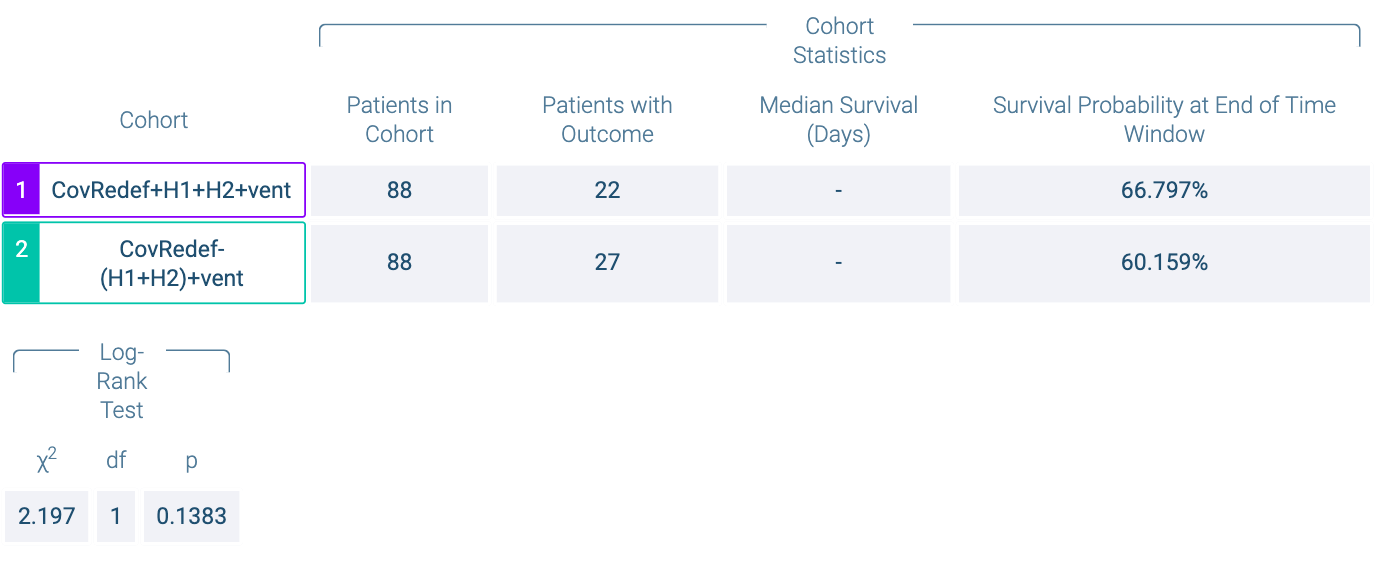

### Outcome_1_Result_b_KM_table.png

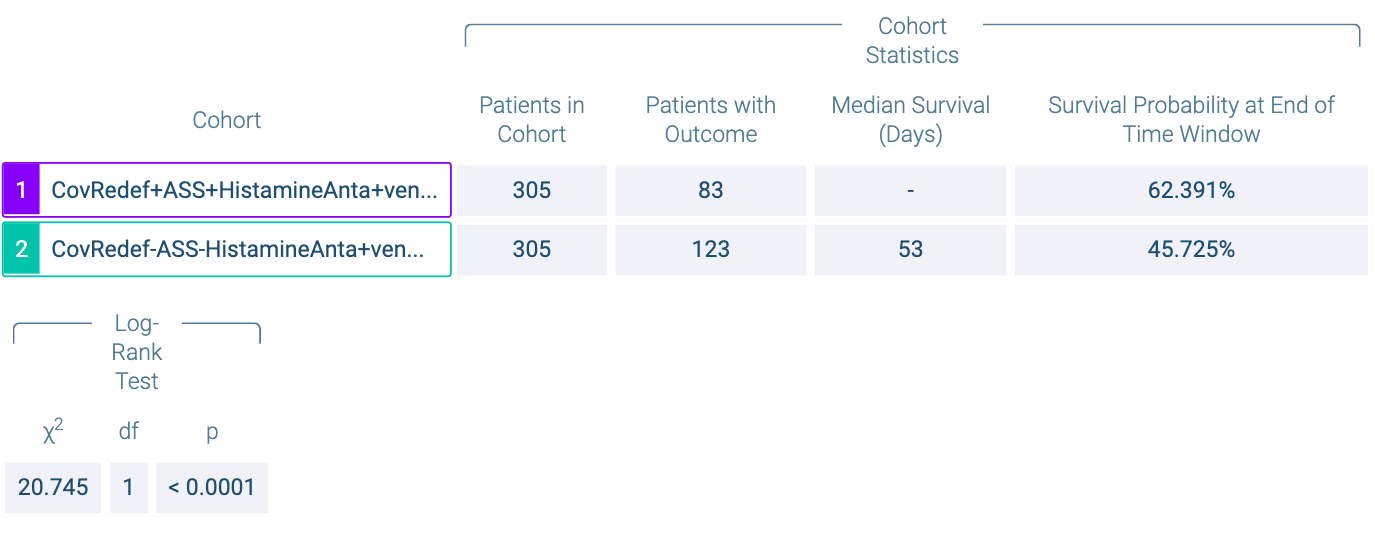

### Propensity_Score_Density_Graph_Large.png

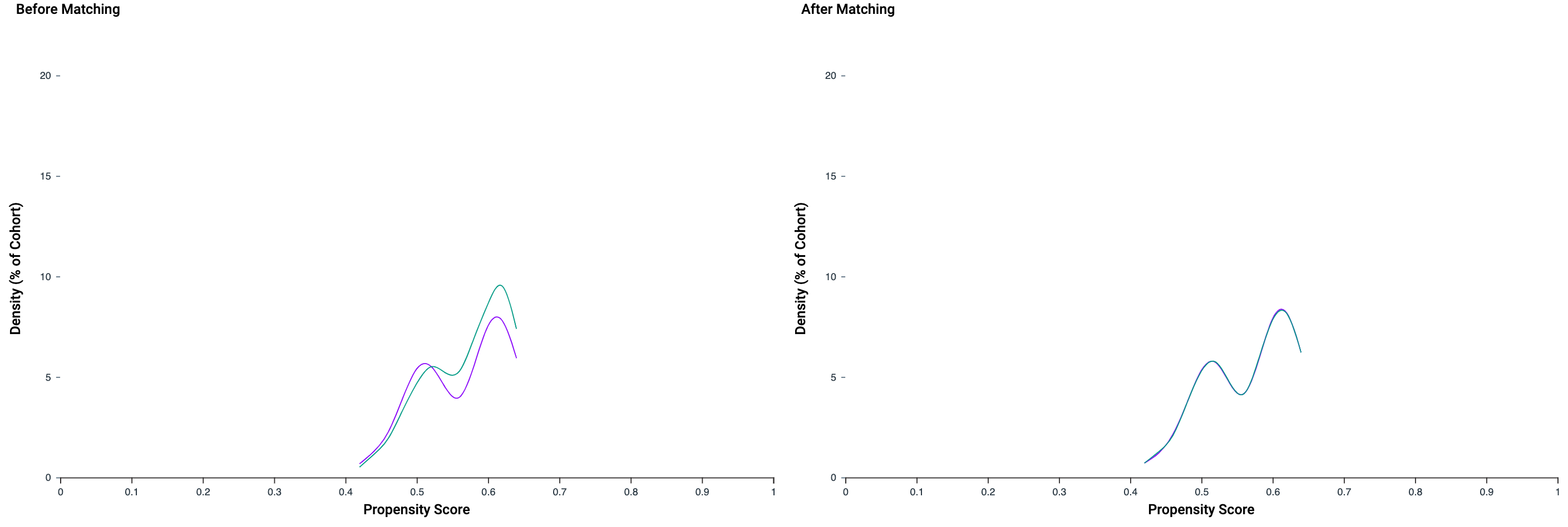

### Propensity_Score_Density_Graph_Large.png

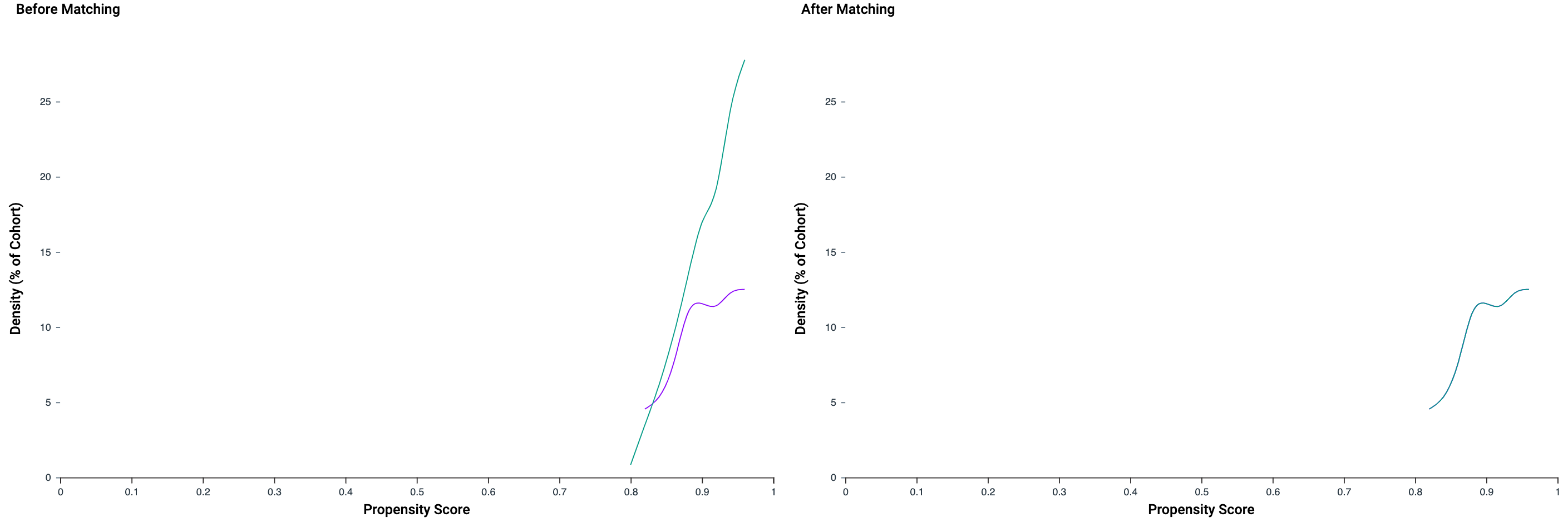

### Propensity_Score_Density_Graph_Large.png

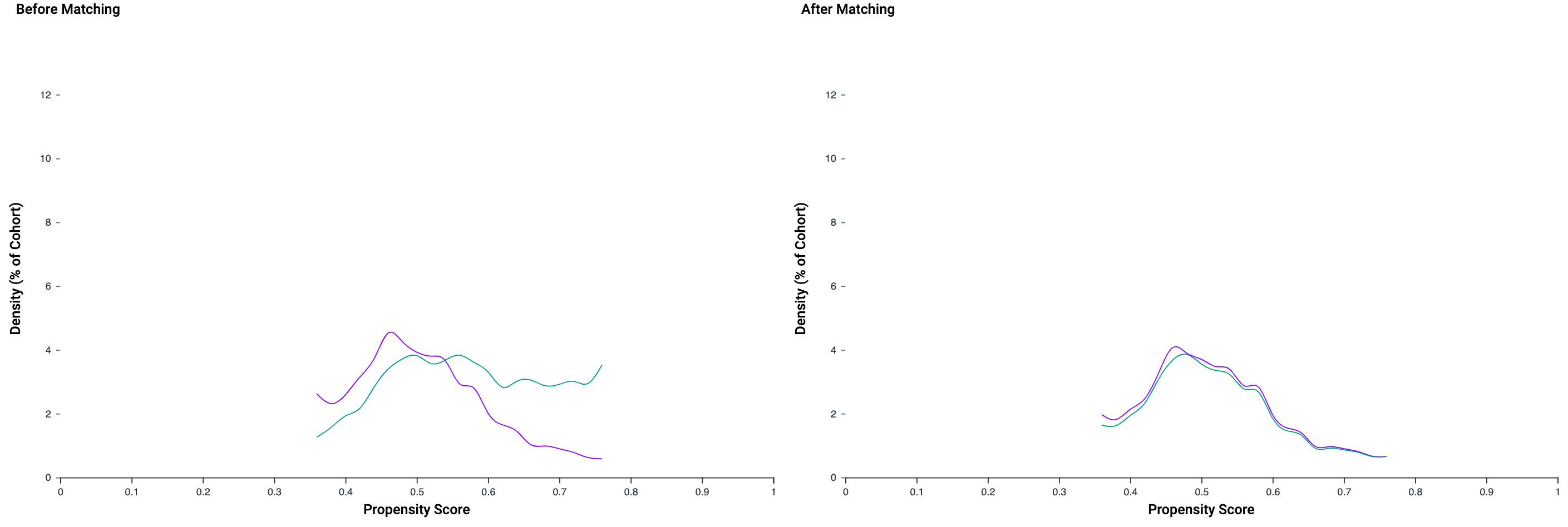

### Propensity_Score_Density_Graph_Small.png

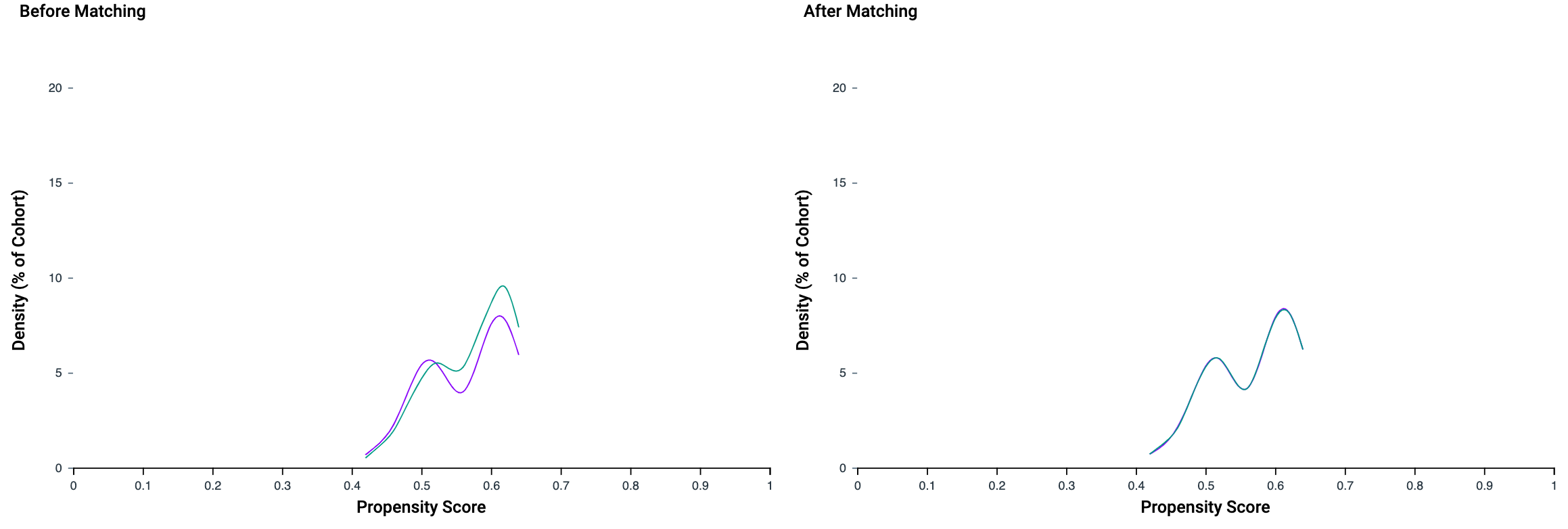

### Propensity_Score_Density_Graph_Small.png

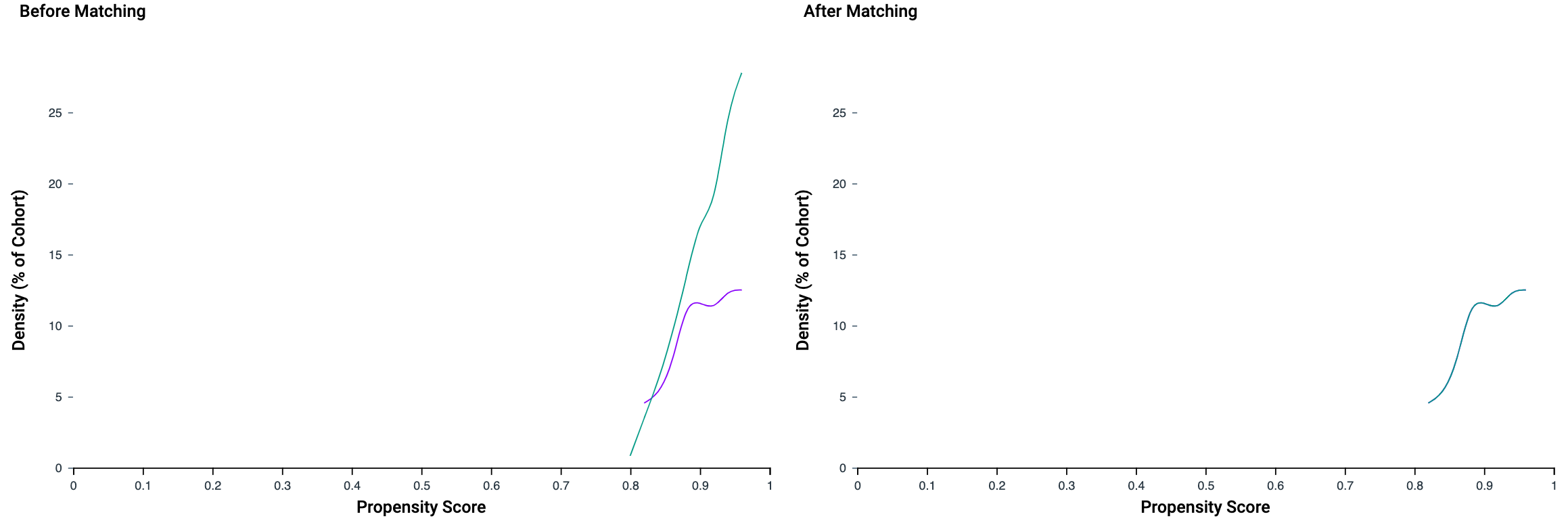

### Propensity_Score_Density_Graph_Small.png

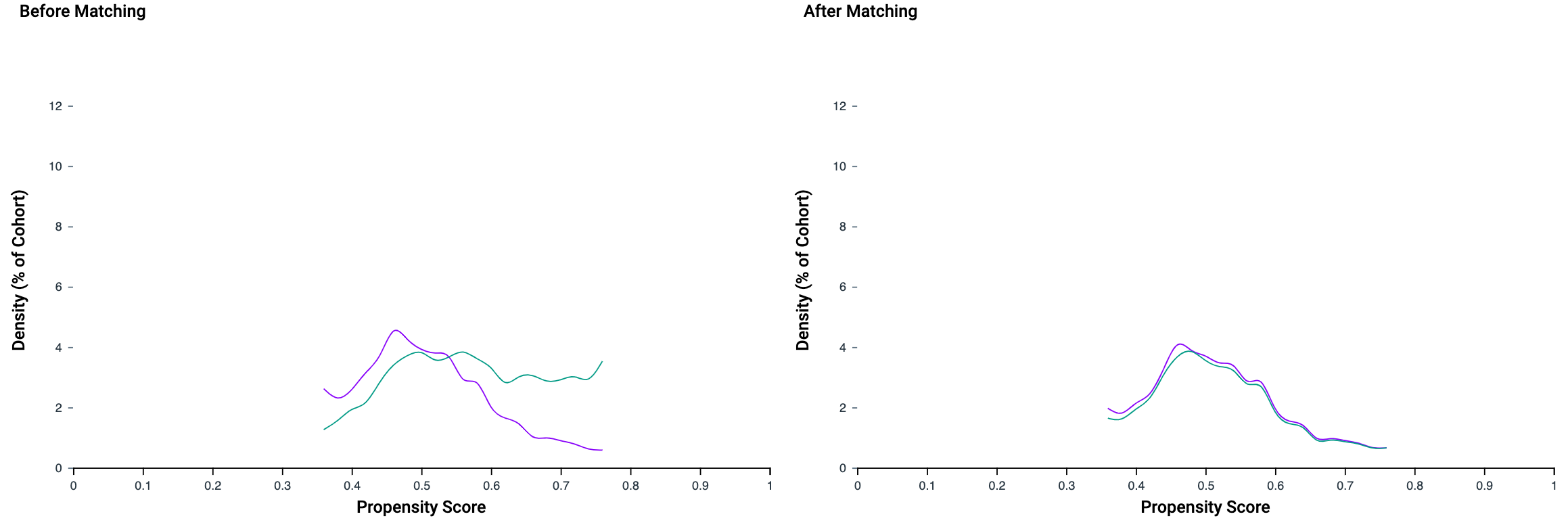
